# A scoping review of the unmet needs of patients diagnosed with idiopathic pulmonary fibrosis (IPF)

**DOI:** 10.1101/2023.09.11.23294619

**Authors:** Carita Bramhill, Donna Langan, Helen Mulryan, Jessica Eustace-Cook, Anne-Marie Russell, Anne-Marie Brady

## Abstract

**Title:** A scoping review of the unmet needs of patients diagnosed with idiopathic pulmonary fibrosis (IPF).

**Aims:** Patients diagnosed with IPF have a high symptom burden and numerous needs that remain largely unaddressed despite advances in available treatment options. There is a need to comprehensively identify patients’ needs and create opportunities to address them. This scoping review aimed to synthesise the available evidence and identify gaps in the literature regarding the unmet needs of patients diagnosed with IPF.

**Methods:** The protocol for the review was registered with Open Science Framework (DOI 10.17605/OSF.IO/SY4KM). A systematic search was performed in March 2022, in CINAHL, MEDLINE, Embase, PsyhcoInfo, Web of Science Core Collection and ASSIA Applied Social Science Index. A comprehensive review of grey literature was also completed. Inclusion criteria included patients diagnosed with IPF or PF; English language only and date range 2011-2022. A range of review types were included. Data was extracted using a data extraction form. Data was analysed using descriptive and thematic analysis. A total of 884 citations were reviewed. Ethical approval was not required.

**Results:** 52 citations were selected for final inclusion. Five themes were identified: psychological impact of an IPF diagnosis; adequate information and education: at the right time and in the right way; high symptom burden support needs; referral to palliative care and advanced care planning (ACP) and health service provision-a systems approach.

**Conclusion:** This review highlights the myriad of needs patients with IPF have and highlights the urgent need for a systems approach to care, underpinned by an appropriately resourced multi-disciplinary team. The range of needs experienced by patients with IPF are broad and varied and require a holistic approach to care including targeted research, coupled with the continuing development of patient-focused services and development of a clinical care programme.

## 1 Introduction

### 1.1 Background

Interstitial lung disease (ILD) describes a range of heterogeneous lung conditions characterised by inflammation and fibrosis of the lung interstitium. (1, 2) A large proportion of patients diagnosed with ILD have pulmonary fibrosis (PF), most commonly Idiopathic pulmonary fibrosis (IPF), representing around 17-37% of all ILDs. (3) IPF is a chronic progressive and irreversible disease which can profoundly and devastatingly impact the physical and psychological well-being of individuals. (4, 5)

People living with IPF may experience debilitating symptoms, which vary in severity and disease course. Symptoms include, progressive worsening breathlessness, impaired lung function, cough, fatigue, (6, 7) with many patients and their carers experiencing anxiety and or depression. (8, 9) This high symptom burden, (1, 10) coupled with social isolation for some, an inability to perform daily activities and the adjustment to a reduced life expectancy (median survival being 2-5 years) can impact quality-of-life (QoL). (11)

Affecting predominantly older adults (12) the incidence of IPF increases with age and with higher rates seen in males. (13, 14) Internationally, there has been a lack of standardisation in diagnostic coding leading to an estimated reported prevalence of IPF ranging from 7 to 1650 per 1001000 persons. (15)

Incongruence persists between the needs of patients with IPF and the actual delivery of healthcare services to adequately meet their needs. Individual needs of patients with IPF are important and so a person-centred approach encompassing the multiple components of the wider healthcare delivery system is needed. Addressing unmet needs for patients with IPF will contribute to improved QoL. (16, 17) Several studies including systematic reviews investigated the care needs and experiences of patients with PF. (5, 6, 7, 16, 17, 18, 19, 20, 21) Many pre- COVID-19 studies have a broadly hospital-based focus with minimal recognition of the changing landscape of healthcare delivery including community-based care.

Informed decision-making is integral to IPF pathways of care and where this is not facilitated, further unmet needs in healthcare and patient preferences may result. Despite advancements in our understanding of the pathogenesis of the disease and the ongoing delivery of antifibrotic treatment, deficits in our understanding of the needs and research priorities of patients with IPF/PF prevail.

### 1.2 Aim

This scoping literature review aims to examine the unmet needs of patients living with a diagnosis of IPF.

### 1.3 Objectives

1. To synthesise the unmet needs of patients living with a diagnosis of IPF.
2. Define barriers and facilitators to meeting patient’s needs.
3. Provide an overview of relevant concepts and terminology.

### 1.4 Registration

The protocol for this study has been registered at Open Science Framework with its unique identifying number DOI 10.17605/OSF.IO/SY4KM

## 2 Methods

### 2.1 Eligibility criteria

We utilised the PCC framework (population (patients with IPF), concept (unmet needs), and context (ILD specialist clinical services) to define the search strategy inclusion criteria. (22) Studies describing the unmet needs of patients living with a diagnosis of IPF were included. We included sources that related to patients with a diagnosis of PF, where IPF was included, given the similarities in the disease course. The search was limited to human participants >18 years of age at the time of data collection (PCC Framework for search strategy development is available in appendix 1).

### 2.2 Scoping Review of the evidence

This review was conducted in accordance with the Joanna Briggs Institute framework for scoping reviews and included the following steps: (i)identifying the research question, (ii)developing a search strategy, (iii)study selection and (iv)data analysis and presentation. (22) The Preferred Reporting Items for Systematic Reviews and Meta-Analyses (PRISMA-ScR) checklist guides the reporting quality of this review. (23)

### 2.3 Sources & searching

Europe. The search strategy was developed with the assistance of a medical research librarian (JEC) and externally peer-reviewed by a second librarian as per the Peer Review of Electronic Search Strategies (PRESS) guidelines. (24) Six databases were systematically searched including, Medline, CINAHL, APA PsycINFO, (EBSCO platform), Embase (Elsevier), and Web of Science (Core Collection) and ASSIA (Applied Social Science Index) (Proquest), on 14^th^ November 2022 and updated on 12^th^ January 2023 to identify studies that met the review’s inclusion criteria. Date limit criteria was applied at full text review (2011-present). A restriction to literature published pre-2011 was applied as this was the year that antifibrotic treatment for patients with IPF became available in the UK and Europe.

No language or geographic limits were applied. Grey literature and unpublished studies were included; sources included ProQuest Dissertations and Thesis Global, Google Scholar and ClinicalTrials.gov, WHO International, Clinical Trials Registry Platform and OpenGrey. Further, a comprehensive online search of key websites, a manual search of the reference lists of included studies and a search of annual conference abstracts sought to identify studies that may have been missed within the initial search.

The search strategy and database search were both conceptualised by researcher (CB) and an information specialist (JEC). Key search terms related to ‘IPF’, ‘unmet needs’ and ‘idiopathic pulmonary fibrosis’ & ‘pulmonary fibrosis’ (The search string is available in appendix two).

### 2.4 Study Selection

Initially all identified records were collated and uploaded onto EndNote X9.3.3 (Clarivate Analytics, Pennsylvania, USA) and duplicates removed. Then all identified citations were transferred into Covidence software (Veritas Health Innovation, Melbourne, Australia) where any remaining duplicates were removed.

The next step was to screen the titles, and abstracts conducted by two independent reviewers (CB and DL) for assessment against the inclusion criteria. Potentially relevant studies which met the inclusion criteria were retrieved in full text and uploaded to Covidence. The full text of selected citations was assessed in detail against the inclusion criteria (PCC inclusion criteria) by the same two independent reviewers. Any disagreements which occurred about the inclusion or exclusion of a paper, were discussed by the reviewers until agreement was found. A third reviewer HM arbitrated when there was disagreement about the inclusion of a paper (n-4). We included all review types (systematic, scoping and narrative reviews) which described the unmet needs of our patient group. This review did not include case reports, protocols, letters or posters, commentaries, and opinion pieces.

### 2.5 Data Extraction

Data was extracted from articles and other evidence sources included in the scoping review by one reviewer (CB), using a data extraction tool developed by the study’s research team in adherence with the review objectives (see appendix two). All studies received verification by another reviewer (AMB & AMR). Several important domains were included in the data capture tool which adhered to the JBI guidelines (see appendix three).

### 2.6 Data Analysis

Data were analyzed, utilizing the Braun and Clarke (2006) framework for reflexive thematic analysis (TA). This is a five-stage process for coding and data analysis, includes an initial step of data familiarisation through deep immersion with the information sources. Next, an iterative process of code development, was supported by analysis software NVivo (NVivo version 12.0). (25) The final steps three to five provided the backdrop to the organisation of codes into themes and fostered a rich in-depth analysis of the data culminating in theme development.

## 3 Results

### 3.1 Summary of the studies

After eliminating duplications, 884 unique citations were identified. Of these, 719 records were excluded after title/abstract screening, leaving 165 records for further assessment. In addition, a further 20 records were identified through reference checks of systematic and narrative reviews. After full text review, 113 records were deemed ineligible and were excluded (figure 1, PRISMA). The primary reason for exclusion was the lack of data on the outcome of interest (n=74). Other common reasons for exclusion were wrong population (n=11) and commentary or opinion piece (n=16). Fifty-two information sources met the inclusion criteria, of which 52% were published in the last five years. All included articles were published between the period of 2011 to 2022. All included studies were published in English and were from a wide geographical area (figure 2). There, was a total of thirty-eight studies included in this review, representing 73 % of included sources, with the remaining 26.9% of sources being a mix of literature reviews (11.5%), reports and guideline type sources, incorporating various type of methods (15.3%) (table 1).

**Figure 1.**
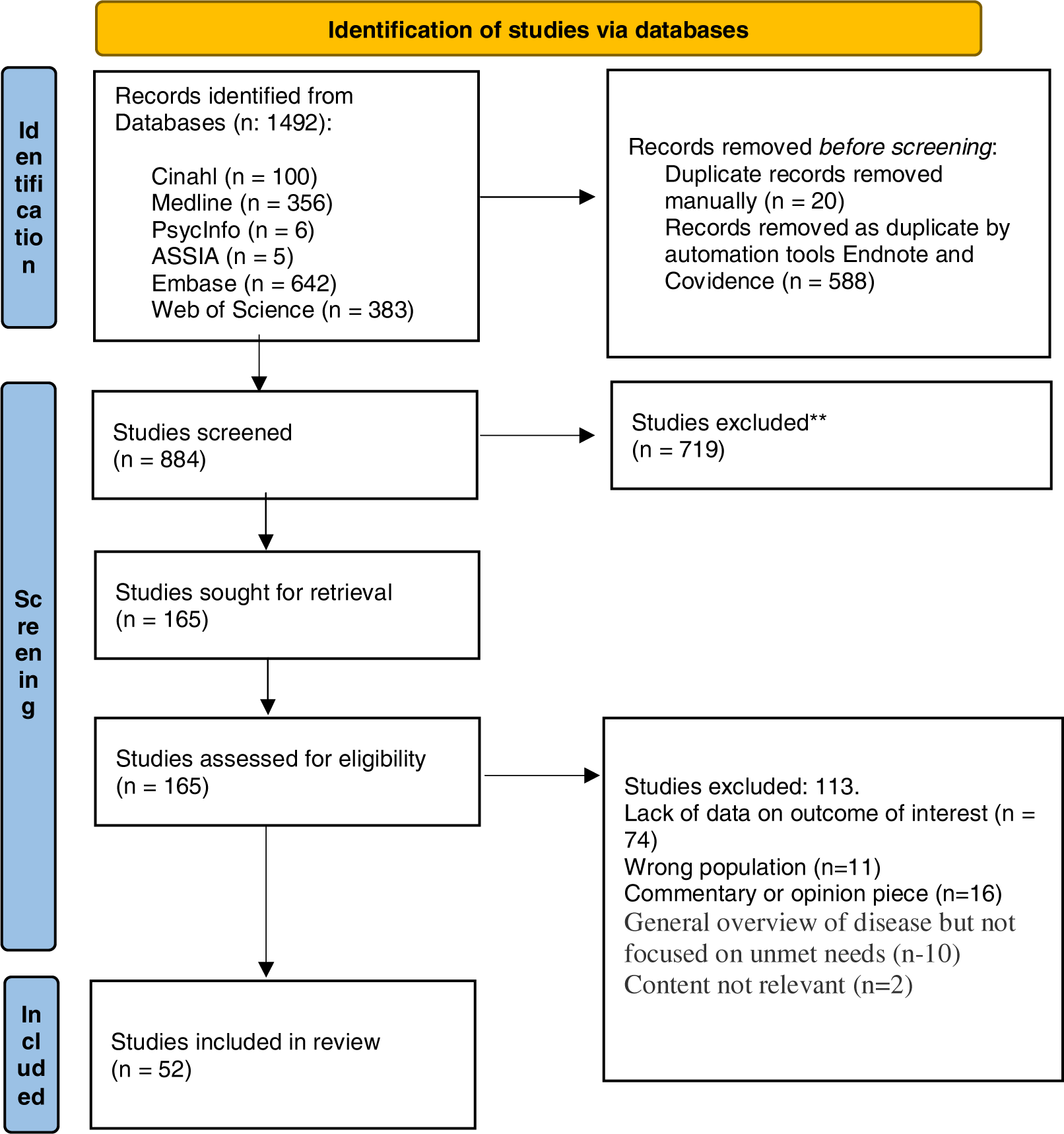
Preferred reporting items for systematic reviews and meta-analyses (PRISMA)

**Figure 2.**
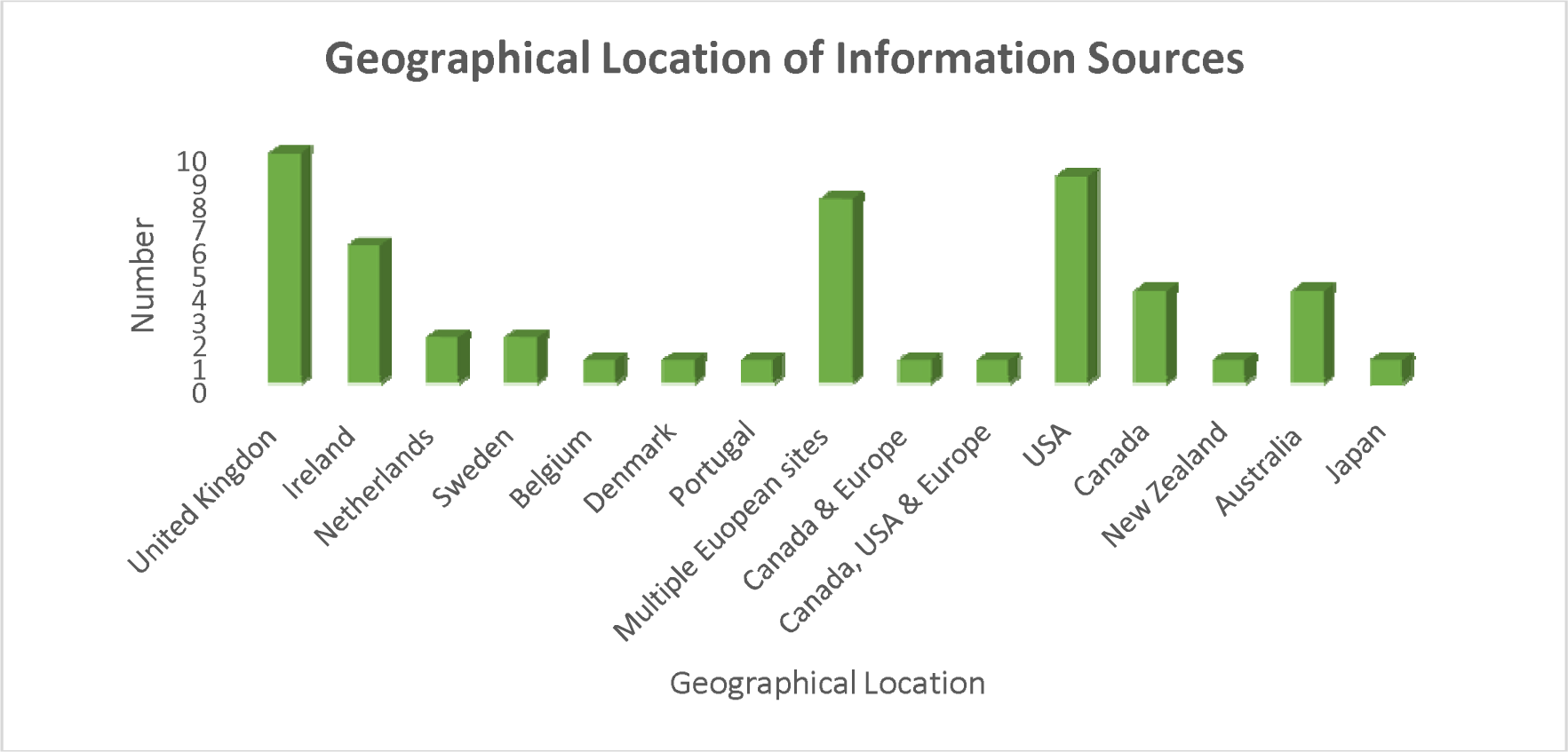
Geographical location of information sources.

**Table 1.** Summary of the characteristics of the included articles (n=52)

| Characteristics | n | % |
| --- | --- | --- |
| **Methodology** | **(N-52)** |  |
| Quantitative | 20 | 38.4% |
| Qualitative | 14 | 26.9% |
| Mixed Methods | 4 | 7.9% |
| RCT | 1 | 1.9% |
| Review | 6 | 11.5% |
| Other: | 8 | 15.3% |
| **Design** | **(N-38)** |  |
| Cross sectional | 25 | 65.7% |
| Cohort | 4 | 10.5% |
| Longitudinal | 7 | 18.4% |
| RCT | 1 | 2.6% |
| **Country** | **(N-52)** |  |
| United Kingdom | 10 | 19.2% |
| Ireland | 6 | 11.5% |
| Sweden | 2 | 3.8% |
| Netherlands | 2 | 3.8% |
| United States | 9 | 17.3% |
| Australia | 4 | 7.6% |
| Canada | 4 | 7.6% |
| New Zealand | 1 | 1.9% |
| Japan | 1 | 1.9% |
| Other European countries | 11 | 21.5% |
| Canda and Europe | 1 | 1.9% |
| USA, Canada and Europe | 1 | 1.9% |
| Multi-site | 23 | 44.2% |
| **Area of interest** | **(N-52)** |  |
| IPF | 40 | 76.9% |
| PF | 4 | 7.6% |
| Fibrotic ILD | 2 | 3.8% |
| ILD | 3 | 5.7% |
| Chronic disease | 1 | 1.9% |
| Respiratory health | 1 | 1.9% |
| Death and dying | 1 | 1.9% |
| **Methods of data collection** | **(N-38)** |  |
| Questionnaire | 13 | 34.2% |
| Semi-structured interview | 9 | 23.6% |
| Focus group | 4 | 10.5% |
| Registry data | 4 | 10.5% |
| World Cafe | 1 | 2.6% |
| Chart review | 2 | 5.2% |
| Mixed methods | 4 | 10.5% |
| RCT | 1 | 2.6% |
| **Outcomes addressed** | **(N-38)** |  |
| Early accurate diagnosis | 18 | 47.3% |
| Palliative care and ACP | 25 | 65.7% |
| Information needs | 27 | 71% |
| Psychological and emotional needs | 20 | 52.6% |
| Care experiences | 12 | 31.5% |
| Treatment pharmaceutical | 25 | 65.7% |
| Geographical location | 6 | 15.7% |
| Multi-disciplinary team | 6 | 15.7% |
| E-health | 4 | 10.5% |
| Lung transplant | 12 | 31.5% |
| Impact on relationships | 8 | 21.05% |
| Access to tertiary centre | 9 | 23.6% |
| Physical burden of IPF | 14 | 36.8% |
| Oxygen needs | 12 | 31.5% |
| Pulmonary rehabilitation | 14 | 36.8% |
| Carer experience | 15 | 39.4% |
| ILD Nurse | 9 | 23.6% |
| Clinical trial access | 3 | 7.8% |
| **Area of interest (non-study sources)** | **(N-14)** |  |
| Pharmaceutical | 1 | 7.1% |
| Palliative care | 3 | 21.4% |
| Care challenges | 4 | 28.5% |
| Respiratory health | 1 | 7.1% |
| Chronic disease | 1 | 7.1% |
| Management of IPF | 4 | 28% |

Of the studies (n=38), 52.6% studies employed quantitative approaches (n=20), with others using a qualitative design, 36.8% (n=14), with further studies employing a mixed method methodology 10.5% (n=4). Several literature reviews (n=6) incorporated a range of methods. The remaining information sources were diverse (n=8) including guideline documents (n=2), patient charters (2), framework documents (n=1) reports (n=2) and a position statement (n=1). Information sources originated from multiple sources.

Of the thirty-eight studies included in the review, >31% of included studies (n-38) investigated care experiences (n=12). A further 66% of studies explored palliative care needs and advanced care planning (n=25). The remaining studies predominantly explored communication/ information needs 71% (n=27), pharmaceutical treatments 66% (n=25), early and accurate diagnosis (n=18), psychological and emotional needs (n=20). Other areas evident were impact on relationships (n=8), physical burden of IPF (n=14), oxygen needs (n=12), pulmonary rehabilitation (n=14), carer experience (n=15), and geographical location (n=6), access to a tertiary centre (n=9), multidisciplinary team (n=6), ILD nurse (n=9), tele-health (n=4), lung transplantation (n=12) and clinical trial access (n=3) (Table 2).

**Table 2.** Unmet needs of patients diagnosed with IPF/PF identified in the included evidence sources.

|  | N | Quantitative | Qualitative | Mixed Methods | Review Report Guide line | RCT |
| --- | --- | --- | --- | --- | --- | --- |
| Carer experience | 13 |  | (26) (20) (27) (28) (12) (29) 30) (31) | (5) (32) | (14) (18) | (33) |
| Pulmonary Rehabilitation | 15 | (17) (4) | (19) (34) (16) (30) (35) (36) | (5) (37) | (11) (38) (39), (38) (40) |  |
| Information needs | 26 | (17), (41), (17) (42) (43),(44) | (28), (30), (45), (27), (29) (46) (47) (16) (34) (35) (44) | (5) (32) (48) | (11) (49) (14) (18) (40) | (33) |
| Psychological/E motional needs | 19 | (42) (17) | (31) (34) (19, 20, 27) (28) (30) (44) (50) (36) | (37) | (11) (49) (14) (18) (40) | (33) |
| Physical burden of IPF | 14 | (51) (52) | (19) (50) (35) (31) | (5) | (11) (14) (18) (49) (53) (39) (38) |  |
| Palliative Care and ACP | 22 | (54) (17) (55) (4) (56) (52) (57) (58) (59) | (28, 34, 47) (20) (6) (16) (30) (36) |  | (11) (49) (60) (40) (39) | (33) |
| Impact on relationships | 7 | (51) | (19) (27) (20) (6) (50) | (5) |  | (33) |
| Oxygen needs | 12 | (58) | (19) (20) (16) (29) 35) (36) | (37) | (11) (49) (14) (40) |  |
| Early accurate diagnosis | 19 | (7), (17), (4) (58) (41) | (20) (34) 50) (6) (16) (35) (44) (19) (36) | (5) | (11) (39) (38) (40) |  |
| Care experiences | 12 | (43) (7) (17) (43) | (35), (34), (20) (61) (27) (31) | (5) | (11) |  |
| Treatment pharmaceutical | 25 | (7) (62) (63) (58) (41) (17) (4) 43) | (34) (35) (16) (27) (30) | (48) (18) (37) | (49) (64) (14) (18) (39) (40) (38) |  |
|  |  | (58) (41) |  |  |  |  |
| Geographical location | 6 | (65) (66) (67) (43) | (28) (35) |  |  |  |
| Multidisciplinary team Discussion | 6 | (17) (58) | (34) (35) | (37) | (39) |  |
| E-health | 4 |  | (12) (32) (34) | (37) |  |  |
| Lung transplant | 11 | (67) (17) (4) | (16) (28) (30) (35) | (37) | (38) (39) (40) |  |
| ILD Nurse | 9 | (17) | (34) (27) | (37) | (11) (49) (38) (39) | (33) |
| Access to a tertiary centre | 9 | (68) (58) (41) (17) (65) (4) (41) | (50) (31) |  |  |  |
| Clinical trial | 3 | (58) | (35) | (32) |  |  |

Recruitment strategies within the literature was reported to be via single sites (n=15) or multiple sites ranging from two to fourteen sites (n=23). A homogenous IPF sample was investigated in most sources (84%). Remaining studies featured heterogenous samples, included other ILD’s, specifically F-ILD (5.2%) and pulmonary fibrosis (8%) and ILD (2.6%).

Male participants were significantly more frequently represented in all included studies (range 41.3%-87.7%); age range of 20-90 years. Studies which reported ethnicity or race of participants included white backgrounds (n=5). Only a limited number of studies included information on employment status (n=2), education (n=4), insurance (n=3) and marital status (n=1).

### 3.2 Reflexive thematic analysis

The process of reflexive thematic analysis enabled the development of five themes relating to the unmet needs of patients living with a diagnosis of IPF (table 3).

**Table 3.** Theme Development.

| Codes | Subthemes | Major themes |
| --- | --- | --- |
| Psychological needs<br>Emotional well-being<br>Social needs | Psychosocial needs.<br>Systemic impact on self and family members.<br>The impact of anxiety, depression & stress.<br>Social isolation.<br>Coping strategies.<br>Emotional distress.<br>Peer support. | **Psychological/Emotional impact of an IPF diagnosis** |
| Information needs<br>Information on living well with IPF | Inadequate information provision.<br>Consideration for timing of gradual and empathetic information about IPF.<br>Patient advocacy groups and their role in information provision.<br>Varied experiences of information and support, particularly at time of diagnosis.<br>Review understanding of disease, trajectories, and impact regularly with the patient. | **Adequate information and education, at the right time and in the right way** |
| • Symptom experience<br>• Functional needs<br>• Physical needs | • Influence of symptoms on well-being.<br>• Symptoms and co-morbidities increase complexity of care.<br>• High symptom burden.<br>• Impact of medication side effects on patients.<br>• Recognition of supportive care needs.<br>• Impact of sexual dysfunction on relationships. | **High symptom burden support needs** |
| • Open and honest conversations about end-of-life care options | • Have earlier open conversations.<br>• Start conversations earlier with a specialist.<br>• Review goals and hopes.<br>• Prepare for end-of-life care with options.<br>• Preferred place of dying.<br>• Discuss options and choice. | **Referral to palliative care and advanced care planning** |
| • Clinical care programme.<br>• Early and accurate diagnosis.<br>• Socio-economic burden on healthcare system. | • Access to anti-fibrotic medication and managing side effects.<br>• Access to clinical trials.<br>• Regular monitoring and care from a multidisciplinary team including an ILD specialist nurse.<br>• Burden of travel to access care.<br>• Appropriate referral for lung transplant assessment.<br>• GP as partner in care.<br>• Access to specialist centres.<br>• National ILD Registry.<br>• Access to E-health.<br>• Access to pulmonary rehabilitation. | **Health service provision - an integrated systems approach** |

### 3.3 The psychological and emotional impact of an IPF diagnosis

A need exists for psychological and emotional support throughout the disease course for patients diagnosed with IPF (table 2). (17, 42) The literature reports that the psycho-social needs of patients (34) and their family carers are frequently being overlooked, (6, 27) with a continued lack of psychological supports for patients diagnosed with IPF. (27) (8) Psychological distress is reported by many patients living with IPF, including worry, fear, anxiety, hopelessness and helplessness. (6, 11, 20, 27) It is reported that many patients with IPF and their carers experience anxiety and or depression. (8, 20, 27, 69)

Patients diagnosed with IPF report a loss of independence coupled with feelings of powerlessness and social isolation (20) The initiation of oxygen therapy is viewed as a particularly stressful time for patients (20), representing for some a distressing trajectory in the disease course and signalled in some cases a loss of hope (27). In one study, oxygen initiation was associated with feelings of shame (34) as the condition became externally visible to others. (27) Patients can also experience increased anxiety related to worry associated with having adequate supplies of prescribed oxygen. (34)

Glaspole and colleagues explored the frequency of prolonged anxiety and depression among people living with IPF, and factors contributing to their persistence. They reported that dyspnoea is a major contributor to anxiety and depression followed by cough, which is also an important contributor. (70) Moor et al. found that although patients were not being specifically asked about access to psychological support in their study, 10% of patients spontaneously reported a need for (better) psychological support throughout the disease course. (17) Van Manen and colleagues highlight the important role an ILD specialist nurse can play in helping patients to manage symptoms such as depression and anxiety particularly as nurses will most likely have been involved in the patients care for some time and may be viewed by patients as someone, they can confide in. (49)

#### Caregiver’s support needs

Significantly the literature reported on the psychological impact of an IPF diagnosis on family caregivers who reported feelings of loneliness and anxiety (20, 27) particularly often associated with the fear of losing their loved one. (45, 69) Caregivers are often not adequately prepared to help their loved one and describe a sense of frustration and helplessness. (69) Ramadurai and colleagues coined the concept “Shrinking world syndrome” in relation to caregivers as IPF progresses they are also at risk of social isolation, loneliness and a restricted lifestyle. (12) (45)

### 3.4 Adequate information and education, at the right time and in the right way

Unmet information needs are prevalent for both patients with IPF and their carers in the presence of a varied disease trajectory (5, 16, 34) The European Patient Charter calls for “comprehensive and high-quality information about IPF including its treatment to be made available to patients”. (*16*). Timely delivery of clinically appropriate information regarding diagnosis and treatment is an important cornerstone in the management of patients with a diagnosis of IPF. (39) For effective communication patients and carers want plain language, honesty and empathy. (30) Furthermore, patients and carers want information with attention to timing, (30) content, (20) and the structure and format of information. (5)

Practical information needs include information on medication use and potential side effects, (30) supplemental oxygen use, nutrition, exercise (61), management of cough and breathlessness (5), insurance cover, travel advice particularly for those on oxygen, trusted online information resources (12) legal and practical advice for disease progression and end-of-life and palliative care planning for patients (16, 54) with F-ILD (28). Patients living alone expressed the most direct and urgent need for information about future care planning (5) There is an emerging need for information on research related the outcomes of clinical trials for IPF. (12, 21) Many patients and carers are not well informed about how their disease will progress. They require information on what to expect and how to prepare for the future (11, 12, 27, 71) with an emphasis on an individualised approach. (50) (12)

It is understood that access to information and education from a diverse range of sources enhances patients and carers coping strategies. (28, 49) Education and reliable information are the bedrock of patient care and help to empower patients to play an active role in their care. (49)

Patients regularly turn to online sources of information on IPF, but these can be of poor quality, outdated or not available in the patient’s native language. (27, 72) Russell and colleagues found that the level of disease awareness varied extensively between patients and reported that approximately one third of patients felt inadequately aware of or informed about IPF. (27)

Caregivers often felt inadequately prepared for the caregiving role and expressed a need for information and education on strategies to help their family member manage IPF and for some they require information on palliative care and advanced care planning (ACP). (28) (27)

Furthermore, there are increased calls for more awareness of PF/IPF among GPs, nurses and physicians. (17) and the general public. (16) Healthcare professionals have expressed a lack of time to discuss the diagnosis and treatment options (60%) with only 39% of HCPs reporting that they had received any training in patient centred communication. (17)

### 3.5 High symptom burden

A significant unmet need related to IPF is the burden associated with the physical and psychological impact of the condition. IPF remains an unpredictable disease of variable course, which could benefit from a systems approach to care, coordinated by a multidisciplinary team. (*5*) The deterioration in health related quality of life for patients with IPF is highly correlated to worsening of symptoms, including worsening breathlessness, cough, and fatigue over time. (11) Lindell et al. analysed focus group data and highlighted that symptoms introduce an overwhelming burden for both the patient and carer, with cough being a particularly challenging symptom. (6) For some patients coughing led to distressing symptoms such as incontinence. (61) Patients report struggling with lethargy which can impact even the simplest of tasks such as reading and watching television. (19)

Several patients in Duck’s study reported feeling depressed which was associated with a lack of control and having to relinquish roles once held. (19) There is growing evidence that daily activities, recreation, pleasure and employment are significantly affected by the burden of symptoms such as anxiety, depression and social isolation connected with a diagnosis of IPF. (11) There were also reported symptom from the side effects of medication in particular antifibrotic treatment adding to the burden of symptoms already experienced by patients (35)

Sampson and colleagues, call for a more pragmatic needs assessment to include components of physical and social functioning, nutrition and symptom burden which would support patients’ self-management and assist with their understanding of the illness and its varied disease trajectories. (5)

### 3.6 Health service provision -a systems approach

Reliance on healthcare services is immense, for those living with IPF with high healthcare costs in terms of resourcing and utilisation of services, provision of multi-disciplinary care and a recognised marked socioeconomic burden for patients. (73) (51) Patients with IPF require regular routine monitoring and input from the multidisciplinary team, including provision for repeated hospitalization. There is a need for increased supportive care particularly at the end-of-life. (51)

A major unmet need in IPF care is the provision of timely and accurate diagnosis (12, 17, 19) and this is a recurrent deficit in health service provision. Several studies have highlighted the scope of the problem with many patients diagnosed with IPF, experiencing a protracted route to diagnosis (17, 34) (6, 16, 35, 44) (19, 36) leading to unnecessary delays including accessing pharmacological interventions and other supportive treatments. (74)

There is an increasing recognition of the importance of having a well-resourced and appropriately staffed multi-disciplinary team (MDT) in providing care to patients with IPF. (37) Current guidelines call for multi-disciplinary discussion involving expert respiratory physicians, radiologists, and pathologists as the gold standard for IPF diagnosis. (1) However, evidence exists which suggests that there can be gaps in staffing some of these MDTs. (37) (75)

Several studies highlight that nonpharmacological treatment options are not equally available for patients in different European countries (16)

Clinical nurse specialists are an essential component of the MDT and are critical to the delivery of holistic care. (19) (39, 76) However there can be variability in access to specialist nursing across jurisdictions. (77, 78) Access to pulmonary rehabilitation (11) can be fragmented with one study reporting that just 42% of patients had access to outpatient pulmonary rehabilitation with similar findings reported for access to psychological support (58%). (17)

From a systems approach to healthcare there is an urgent need for the establishment and recognition of a national registry to capture epidemiologic information. (*39, 62*) There is also a need reported by healthcare professionals to recognize the importance of giving patients the option to participate in research or clinical trials related to IPF. (17, 19) (11)

### 3.7 Referral to Palliative Care and ACP

The World health organisation recommend early palliative care intervention to improve quality of life of patients and their family facing problems associated with life-threatening illness. Since the course of IPF is unpredictable early palliative care interventions can be beneficial in a multiplicity of ways including symptom management, (16, 79) emotional support and in the initiation of advance care planning conversations. (53)

The European Patient IPF Charter and the Irish Thoracic Society Position Statement on the management of IPF have identified an urgent need to involve palliative care in IPF. (16, 28) The National Institute of Health and Care Excellence (NICE) guidance is that patients with IPF should have access to palliative and supportive care services to manage symptoms. (55, 76) Startingly, despite these recommendations, patients with IPF do not receive optimal palliative care over the course of their disease resulting in high symptom burden and decreased quality of life for patients with ILD. (60, 79) There is increasing evidence that patients with IPF do not always have access to palliative care input and when it is introduced the timeline is rarely optimal. (17, 28)There continues to be poor referral and access to palliative care specialists with some healthcare professionals reporting a lack of training in palliative care, (53) insufficient communication training in facilitating these conversations, variations in the disease trajectory (54) and patients preferences to have these conversations. (6, 47, 80) In one study HCPs reported that palliative care was not initiated until the later stages of PF/IPF with a fifth reporting that palliative care was only initiated at end of life. (17) Lack of advance care planning leads to longer ICU and overall hospital lengths of stay. (81) (28)

Kalluri and colleagues in their qualitative study revealed that advance care planning is desired by patients and caregivers early in their illness experience with HCPs citing a need to clarify role, scope and responsibility with a call for practical guidance and training for HCPS to improve competency and confidence in these conversations.

#### Patient and Public Involvement

Patient and public representatives (n= 5) from the Irish Lung Fibrosis Association PPI group were involved in reviewing the research protocol for this scoping review. The stakeholders comprise of patient representatives, family members of patients diagnosed with IPF and experts and researchers in the field of IPF. The scoping review protocol was presented to the group for feedback and discussion during a zoom call meeting. The scoping review findings were presented to healthcare professionals in the field of IPF, in person for further opportunities for discussion. The involvement of PPI in this scoping review has enriched the review and reflected the importance of including those impacted by IPF in the process.

### 3.8 Discussion

In recent years our knowledge and awareness of the complexities and disease burden associated with an IPF diagnosis has improved including the recognition of the diverse range of patient needs. There is increased awareness of the broader impacts of an IPF diagnosis including impacts to patients, family members and the healthcare system.

This scoping review characterised the broad and varied unmet needs of patients living with a diagnosis of IPF and similarly for many patients with progressive PF given the similarities between them. Despite advancements in drug therapies including the use of antifibrotic medication which when introduced heralded improved survival rates for many, there remains multiple unmet needs and compromised quality of life for many patients living with IPF.

The literature revealed that psychological distress was reported by many patients living with IPF and incorporated several elements including anxiety and depression. (6, 20, 27) Psychological distress was heightened particularly in terms of delays in diagnosis and represents a critical issue as regards optimal care pathways for patients with IPF. (34)

Patients have specific requirements regarding the delivery of appropriate and bespoke information at specific time points during the disease trajectory, which remain largely unmet. Furthermore, the need for improved awareness of IPF and tailored information amongst the public and healthcare professionals remains.

Patients living with IPF rely heavily on the healthcare system representing a significant need for an integrated health systems approach to providing supportive care. The literature reported that there are further unmet needs for patients with IPF, including the need for appropriately resourced multidisciplinary teams including the requirement for clinical nurse specialists. Furthermore, a need exists regarding access to services and appropriate referral pathways such as lung transplant assessment referral, (82) psychological support, pulmonary rehabilitation, palliative care referrals and advanced care planning and opportunities to participate in clinical trials. Many of these needs were previously highlighted in a European wide survey on gaps in care for patient living with PF, (17) representing a critical area of focus for future research and funding.

It is essential that we begin to recognise the importance of palliative care as part of the overall management plan and care package for patients with IPF. There are further unmet needs regarding healthcare systems, having established and resourced patient registries to accurately capture epidemiological data including incidence and prevalence of IPF (17) and furthermore offering opportunities for prospective follow up of patients. (39)

In summary, the findings of this review extend our current understanding of the broad scope of unmet needs for patients with IPF and highlight the diverse and nuanced approach to care which is required for this patient population.

### 3.9 Study Limitations

The core limitations of this scoping review involve the diverse and varying quality of available evidence with only on RCT included. Similarly mixed methods studies were underrepresented in the review comprising just 7.9% of the overall included resources. Furthermore, there were limited longitudinal studies with just 18.4% represented in this review. The majority of studies focused on patients with IPF (76.9%). The methodological quality of studies was high, yet many excluded some important demographic variables including stage of disease, oxygen use, socioeconomic status, and employment status. Given the varied disease trajectory, the interpretation of the findings from some studies may have been impacted. This review included non-English language articles but cannot claim to have exhausted all non-English resources despite utilising a systematic approach to the review.

#### Conclusion

In recent years we have seen significant advances in relation to our understanding of the pathogenesis of IPF coupled with the introduction of antifibrotics and their recognised contribution to patient survival. The concept of unmet needs and quality of life are intrinsically linked and yet there remains deficits in the literature as regards comprehensively investigating this relationship for patients with IPF representing a need for future research focus examining this relationship.

This review will extend the knowledge base of the muti-disciplinary team as regards the diverse range of needs that patients with IPF and their carers have and signals the need to continue to target research toward this underrepresented patient population. The literature highlights the continued lack of integrated clinical care programmes in many jurisdictions for the management of IPF, which can result in unstructured and fragmented care delivery for patients. This scoping review highlights that patients living with a diagnosis of IPF experience a myriad of unmet needs across a broad range of areas and require a comprehensive multi-disciplinary approach to care with equal access to services and tailored information to support them over the course of the disease and represents a future key area for research.

## Data Availability

All data produced in the present study are available upon reasonable request to the authors

## Contributions

CB and A-MB led the conceptualization, design, and development of this study. DL, HM and AMR assisted with the scoping review approach. CB and J-EC developed the search strategy with input from A-MB. Titles, and abstracts were screened by two independent reviewers (CB and DL) for assessment against the inclusion criteria with HM acting as arbitrator where required. Data extraction was completed by one reviewer (CB). All studies received verification by another reviewer (AMB & AMR). CB drafted the scoping review findings. AMR provided feedback from a clinical perspective on the scoping review manuscript. All authors reviewed and approved the final version of the manuscript.

## Competing interests

None declared.

## Funding

This work was supported by the Irish Research Council; Grant number: GOIPG/2022/56

## Acknowledgement Section

With special thanks to the participants of the Irish Lung Fibrosis Association PPI group.

## Notes

### Competing Interest Statement

The authors have declared no competing interest.

### Clinical Protocols

https://bmjopen.bmj.com/content/13/5/e070513

### Funding Statement

Funding: CB received a PhD scholarship from the Irish Research Council which supports this work; Grant number: GOIPG/2022/56. The Irish Research Council played no role in the preparation of this manuscript. Funding | Irish Research Council

