## Supplementary material for "A scoping review of the unmet needs of patients diagnosed with idiopathic pulmonary fibrosis (IPF)": Appendix 1-4 for Medrxiv.docx

**PCC Framework for search strategy development**

| **Framework** | **Element** | **Key Terms** |
| --- | --- | --- |
| **PCC** | **Population** | Adult patients over the age of 18 years who have a diagnosis of idiopathic pulmonary fibrosis. It will also include articles that refer to patients with a diagnosis of pulmonary fibrosis. |
|  | **Concept** | Healthcare needs. |
|  | **Context** | All care settings. |

**Appendix 2:**

**Search of CINAHL (EBSCO) conducted on 14^th^ November 2022.**

| **Search** | **Query** | **Records Retrieved** |
| --- | --- | --- |
| S1 | (MH "Idiopathic Pulmonary Fibrosis") OR (MH "Idiopathic Interstitial Pneumonias+") OR (MH "Pulmonary Fibrosis+") | 4,017 |
| S2 | TI ( “Idiopathic pulmonary fibros*” OR “Idiopathic interstitial pneumonia*” OR “Familial Idiopathic Pulmonary Fibrosis*” OR “Usual Interstitial Pneumon*” OR “fibrosing interstitial lung disease” OR “progressive fibrosis” OR “nonspecific interstitial pneumonia” OR “pulmonary fibros*” ) OR AB ( “Idiopathic pulmonary fibros*” OR “Idiopathic interstitial pneumonia*” OR “Familial Idiopathic Pulmonary Fibrosis*” OR “Usual Interstitial Pneumon*” OR “fibrosing interstitial lung disease” OR “progressive fibrosis” OR “nonspecific interstitial pneumonia” OR “pulmonary fibros*” ) | 4,333 |
| S3 | TI ((service* OR need* OR support* OR care* OR caring OR nurs* OR pathway*) N4 (access* OR barrier* OR disparit* OR demand* OR gap | 234,910 |
| S4 | (MH "Healthcare Disparities") | 16,029 |
| S5 | (MH "Health Services Accessibility+") OR (MH "Health Services Needs and Demand+") | 120,306 |
| S6 | S3 OR S4 OR S5 | 327,920 |
| S7 | S1 OR S2 | 5698 |
| S8 | S6 AND S7 | 100 |

**Appendix 3: Data Extraction Instrument**

| **Evidence source details and characteristics** |
| --- |
| Author |
| Year |
| Title |
| Country |
| Aims/Purpose |
| Study Population and sample size |
| Concept |
| Context or setting |
| Study Design |
| Methodology |
| Patient needs identified (physical, psychological, other) |
| PROMS Utilised |
| Summary of key findings |
| Limitations |

**Appendix 4: Scoping review sources.**

| **Reference**  **Country** | **Aim** | **Sample size**  **Recruitment** | **Participants** | **Setting** | **% Male** | **Age range (mean) in years** | **ILD Type** | **Methodology** | **Summary of key findings** |
| --- | --- | --- | --- | --- | --- | --- | --- | --- | --- |
| **Quantitative** | | | | | | | | | |
| 1) Akiyama  2020  Japan | This study aimed to characterize the practice of pulmonologists regarding palliative care and end-of-life communication for patients with IPF and identify perceived difficulties and barriers to this. | 135  Multi-site | HCP: Representative sample of pulmonologists from Shizuoka prefecture, Japan. | Pulmonologists working at up to 15 general hospitals in the region. | 87.7% (total sample) | Age range between 20 and 60 with highest percentage (41.5%) in their 30’s | IPF | Quantitative study utilising self-administered questionnaires. | Findings reported that patients with IPF complained of dyspnea and cough. However, less morphine was prescribed for IPF then for lung cancer.  Participants experienced greater difficulty in providing palliative care for IPF patients then for lung cancer patients.  End of life discussions in patients with IPF were conducted later than the physician-perceived ideal timing. |
| 2) Brereton  2020  UK & Ireland | The objective of this study was to investigate, within primary care healthcare systems, the time taken from the first primary care physician referral for symptom investigation to ILD specialist centre review and antifibrotic therapy commencement. | 247  Multi-site | Patients diagnosed with IPF | Recruitment took place at two specialist ILD clinics in two countries with a primary care healthcare system (University Hospital Southampton, Southampton, UK, and Mater Misericordiae University Hospital, Dublin, Ireland) | 74% (total sample) | Patients seen in 12 months or less (n-142) mean age 72 mean SD + 8.8 years.  Patients seen in 12-24 months (n-41) mean age 73.5 SD + 7.39 years; patients seen after 24 months (n-63) mean age 73.3 SD + 6.6 years. | IPF (N=247), | Quantitative study using electronic case note review. | Patients reviewed in the ILD clinic within 12 months of primary care referral had a significantly preserved mean forced vital capacity (FVC) (83±22% predicted) compared to those reviewed between 12 and 24 months (75±22%, p=0.036), and >24 months (71±17%, p⩽0.001), respectively.  Kaplan–Meier analysis identified that patients seen in the ILD clinic within 12 months from the initial primary care referral had significantly longer time to discontinuation of therapy compared to those seen at 12–24 or >24 months. |
| 3)Cove  2015  UK | The aim of this pilot study was to characterise the psychological needs of both patients and carers and to test the feasibility of a workshop approach. | 29  Single site | 13 patients with IPF  16 family members | Workshop | Male Patients: (n-10)  Male Family members: (n-2) | unknown | Patients with IPF (n-13) | Quantitative questionnaire post workshop attendance | Preliminary data suggests attendance at a workshop addressing psychological issues is feasible and is perceived to be a positive experience by patients diagnosed with IPF and their families. |
| 4) Dedent  2021  United States | Assess the geographic differences in mortality and baseline pulmonary function in participants with IPF. | 849  Single site | Patients diagnosed with IPF. | UCSF ILD Clinic | Unknown | Unknown | IPF  (n-849) | Retrospective cohort study  Online data capture system. | Rural participants have decreased baseline pulmonary function and increased mortality within their first year of presentation to a tertiary care centre compared to suburban participants, independent of individual-level characteristics. |
| 5)Johansson  2022  Canada | To test the hypothesis that greater travel distance to access ILD clinical care would be associated with more severe disease at time of referral and worse clinical outcomes. | 1162  Multi-site | Patients with fibrotic Interstitial lung disease | Canadian Registry for Pulmonary Fibrosis (CARE-PF) which is a multicentre prospective registry of patients from ILD subspecialty clinics across Canada | 49%  (Total sample) | Mean age 63 ±12 | Patients with fibrotic ILD (n=1162) | Quantitative study  Online data capture system using registry data. | Patients with fibrotic ILD with a longer travel distance to their ILD clinic had better prognostic indices at baseline but had a higher risk of death or lung transplant in the total cohort and in patients with connective tissue disease related ILD. |
| 6)Lancaster  2021  United states  Japan  France  Germany. | To gain physician and patient perspectives on the pathway to care from symptom recognition to diagnosis and disease burden. | Physicians (n=244) reported data on  Patients with IPF (n=1249)  Multi-site | Healthcare professionals reporting on patients diagnosed with IPF plus a number of patients with IPF also self-reported (n=739) | Adelphi IPF II Disease Specific Programme, with the following countries included France, Germany, Japan and the United States. | 68% (total sample) | Mean (SD) age at diagnosis of 65.6 years (SD 10.61) | IPF (1249) | Quantitative point-in-time study using an online survey. | Misdiagnosis and delayed diagnosis were common in this study; examples included a diagnostic delay of 0.8 years in Germany to 2.0 years in Japan after symptom onset was reported.  In all countries, patients more often reported symptoms in the survey than did their physicians.  On average, patients underwent 7–10 clinical tests before diagnosis.  Patients reported low quality of life with only 50% of patient with moderate/severe IPF were satisfied with their treatment. |
| 7)Lamas  2011  USA | To examine the association between delayed access to subspeciality care and survival time in Idiopathic Pulmonary Fibrosis. | 129  Single centre | Patients who met American Thoracic Society criteria for idiopathic pulmonary fibrosis | New York Presbyterian/Columbia University Medical Center (New York, NY) | 76% | Mean age 63 years | IPF | Prospective cohort study using a questionnaire | The results of this single-centre prospective study demonstrate that delayed access to a tertiary care centre is associated with a higher rate of death from IPF independent of disease severity. |
| 8)Lindell  2021  USA | To determine the feasibility, acceptability and efficacy of a nurse-led early palliative care intervention entitled “A Program of SUPPORT”, in patients with IPF and their caregivers. | 76 dyads  Single centre | Patients with IPF (diagnosed in the year previous to their initial center visit)  Caregiver | University of Pittsburgh Dorothy P. and Richard P. Simmons Center for Interstitial Lung Disease at University of Pittsburgh Medical Center. | Patients’ intervention group:  males, 80%  Control group: males  85% | Patient group  Age 65 (median age of 70 in intervention arm vs. 73 in control arm) | IPF and caregivers (n=76) | A randomized controlled trial . | Efficacy demonstrated a significant improvement in caregiver’s knowledge, disease preparedness, and confidence in caring for the patient as well as an improvement in knowledge and advance care planning completion in patient participants. |
| 9)Maher  2017  United Kingdom | To investigate treatment patterns of European patients with IPF to understand antifibrotic prescribing and identify unmet needs in IPF treatment practice. | 290  Multi-site | Respiratory physicians. | The questionnaire was developed by Elma Research, an independent market research agency, on behalf of F. Hoffmann-La Roche Ltd and distributed to physicians from France, Germany, Italy, Spain, and the UK. | Unknown | Unknown | IPF | Quantitative Study.  Patient chart review followed by an online questionnaire (35-40 min). | This audit highlights the high proportion of patient’s diagnosed with IPF, who do not receive approved antifibrotic treatment.  Overall, 54% of patients with IPF did not receive treatment with an approved antifibrotic. More patients had a confirmed IPF diagnosis in the treated (84%) versus the untreated (51%) population. Of patients with a confirmed diagnosis, 40% did not receive treatment. The treated population was younger than the untreated population (67 vs 70 years, respectively; p ≤ 0.01), with more frequent multidisciplinary team evaluation (83% vs 57%, respectively; p ≤ 0.01). A higher proportion of untreated patients had forced vital capacity > 80% at diagnosis versus treated patients. Of patients with ‘mild’ IPF, 71% did not receive an approved antifibrotic versus 41% and 60% of patients with ‘moderate’ and ‘severe’ IPF, respectively. |
| 10)Moor  2019  Denmark, Ireland, UK, Bulgaria, Czech Republic, Poland, Greece, Italy, Spain, Austria, Belgium, France, Germany and the Netherlands | The aim of the development of the joint expert and patient statement was to highlight the most pressing common unmet needs of patients with PF/IPF. Putting forward recommendations to improve the quality of life and health outcomes throughout the patient journey. | Patients (n=286) and HCP’s (n=69)  Multi-site | Patient survey completed by: Patients 79%, Caregivers 21%  HCP 69:  56 physicians (81%) and 13 specialist nurses (19%). | Online survey distribution by the European Idiopathic Pulmonary Fibrosis and Related Disorders Federation (EU-IPFF) in 14 European countries. | 70 % of the total patient group | Mean age 66 years of the patient group | Patients diagnosed with IPF (n=86%)  Patients diagnosed with another type of PF (n=14%) | Quantitative study using online surveys. | Delays in diagnosis and timely access to interstitial lung disease specialist and pharmacological treatment were identified as important gaps in care.  Patients and HCPs reported that a greater focus on symptom-centred management, adequate information including trial information and increasing awareness of PF/IPF is required. |
| 11)Moor  2021  Netherlands | The aim of this study was to evaluate the predictive value of the surprise question for 1-year mortality in IPF. | Questions answered by 140 HCPS in relation to 140 patients.  Single site | Patients with IPF (n=140) | A prospective cohort study at the Erasmus Medical Center. | 87% (Total population) | Mean age 74 ±6.5 | IPF (n=140) | Quantitative study  Prospective cohort study using a paper questionnaire | The surprise question can accurately predict 1-year mortality in IPF using a multivariable model (OR 3.69; 95% CI 1.24–11.02; *p* = 0.019). The C-statistic of the surprise question to predict mortality was 0.75 (95% CI 0.66–0.85). |
| 12)NCUBE  2020  New Zealand | To create a nurse-led clinic to support, guide and optimise therapy for patients with IPF. | Patients with IPF (n-9)  Single site | Patients diagnosed with IPF. | Single site nurse led clinic. | Unknown | Unknown | IPF (n=9) | Creation of a nurse led clinic: including a quantitative pre/post clinic questionnaire | Needs amongst this small group varied from one patient who was close to end of life and another patient who had just been diagnosed. Patient feedback at initial analysis was positive. |
| 13)Personen  2018  Sweden | This cohort study was performed to explore potential differences in the care of IPF in two Nordic countries. | 312  Finish registry (n-152)  Swedish registry (n-160)  Multi-site | Finnish IPF Registry (n=152)  Swedish IPF registry (n=160) | Two national registries. | Finnish registry mean age (SD)  74.6 (8.3)  Swedish registry 72.5 (8.0) | Finnish registry 68.4%  Swedish registry 70.6% | IPF (n=312) | Quantitative study utilizing two patient registries using online data capture tool. | This study highlighted that there are differences in how patients are treated with antifibrotic drugs in Finland and Sweden.  To be resident in Sweden was the main determinant for receiving antifibrotic drugs (OR 5.48, 95%CI 2.65-11.33, P< 0.0001). |
| 14)Sharp  2017  United Kingdom | To investigate the current practices around palliative and supportive care and to explore the impact of a supportive care decision aid tool. | 73  Single site | The decision aid was prospectively assessed with 73 patients | UK ILD centre. | Pre-tool: 80.9%  Deceased: 82.8%  After-tool: 78.1% | Mean age Pre-tool: 75  Deceased: 75.4  After-tool: 75 | IPF | Quantitative study using a data capture tool. | The decision aid tool was completed for 49.3% of patients and resulted in significant increases in documented discussion of referral to palliative care (11.2% vs 53.6%, p<0.01).  Tool completion led to an increase in referral for palliative care (2.7% vs 16.7%, p<0.01). |
| 15)Swaminathan  2022  USA | To identify clinical characteristics and social determinants of health that differentially associate with lung transplant compared with death in patients with IPF. | 955  Multi-site | Patients diagnosed with IPF | IPF-PRO Registry (A multi-centre US registry of patients with IPF) | Male 75.5% (total sample) | Mean age 70 (65-75 | IPF (N=955 total sample) | Quantitative study evaluating registry data. | For patients on the IPF-PRO Registry, median Zip code, income and access to a lung transplant centre differentially impact the risk of lung transplantation compared with death, irrespective of disease severity measures or other transplant eligibility factors. |
| 16)Turnpenny  2015  UK | This study aimed to explore chest clinicians’ experiences in delivering care in advanced IPF. | 57  Single site | Chest consultants (n=17), chest registrars (n=28), physiotherapists (11), and nurse clinician (n=1). | Questionnaires were distributed at a regional respiratory meeting. | unknown | unknown | IPF | Quantitative study utilising questionnaire. | Less than 10% of all respondents felt they had significant training in initiating End-of-Life discussions or palliating symptoms. |
| 17)Tyas  2019  UK | To explore if the introduction of severity criteria improved palliative care provision for patients with idiopathic pulmonary fibrosis. | 47  Single site | Patients diagnosed with IPF. | Clinic setting | Unknown | Unknown | Clinic letter review of patients with IPF | Quantitative clinical audit tool. | Introducing IPF disease specific markers of severity, following the intervention from a SPCT consultant in 2016, along with having a respiratory consultant with a specialist interest in palliative care, has improved access to palliative care and symptom control for patients with IPF. |
| 18)Van der Sar  2021  Europe | The aim of this study was to evaluate the time to diagnosis of pulmonary fibrosis, identify potential reasons for delays and document patients’ emotions. | 273  Multi-site | Patients with a self-reported diagnosis of pulmonary fibrosis. | EU-IPFF through its member patient organisations in Europe; these organisations distributed the survey to members and other patients through email and social media. | Unknown | Unknown | IPF (n=214)  Sarcoidosis (n=28)  Other types of PF (n=31) | Quantitative study utilizing surveys. | The time to diagnose pulmonary fibrosis varies widely across Europe.  Forty percent of individuals took ≥1 year to receive a final diagnosis.  Delays occur at each stage of the diagnostic pathway. |
| 19)Wysham  2015  Sweden | To compare end of life care in oxygen-dependent Pulmonary Fibrosis patients compared to patients who died from cancer: A national population-based study. | Total: 57,128  patients with PF 285 & patients with terminal cancer 56,843  Multi-site | Patients with pulmonary fibrosis who died from January 1, 2011, through October 14, 2013 &  Patients who died from cancer in the same time period. | Nationwide Swedish Registries of long-term oxygen (LTOT) and EOL care:  Swedevox and Swedish Registry of Palliative Care (SRPC) | PF 61% males  Cancer patients 51% | Patients with PF 78 ±8 Cancer patients 75± 12 | PF (N=285)  Terminal cancer (n=56,843) | Quantitative study utilising nationwide Swedish registries. | PF patients access fewer palliative care services and experience greater symptom burden at the end of life than patients with terminal cancer. |
| 20)Weatherald  2017  Canada | To describe the practice patterns of Canadian respirologists on the diagnosis and management of IPF and to determine if these patterns reflect current guidelines | 112 respondents to the 2009 survey  107 respondents to the 2013 survey.  Multi-site | Canadian Thoracic Society (CTS) members who are academic or community physicians, or respirology fellows-in-training were surveyed. | Canadian Thoracic Society (CTS) member email addresses used for survey distribution. | Unknown | Unknown | The majority of respondents were clinicians (86%) | Online quantitative surveys. | Practice patterns for diagnosis and treatment of IPF in Canada evolved in concordance with the publication of international guidelines in 2011.  Access to palliative care, lung transplantation referral, multidisciplinary diagnosis discussion, and specialty IPF clinics were identified as gaps in IPF care in Canada. |
| **Mixed methods** | | | | | | | | | |
| 21)Maher  2018  Canada, France, Germany, Italy, Spain, United Kingdom | The aim of this study was to investigate the views of patients with IPF and pulmonologists on the diagnosis and management of IPF to understand treatment patterns. | Physician group: (n-287) participated in the online questionnaire and (n-61) in the interview.  Patient group: (n-68) participated in interviews and (n-60) in online questionnaire.  Multi-site | Survey:  Patients with IPF (n=60) Pulmonologists (n=287)  Interview  Patients (n=68) Pulmonologists (n=61) | Patients recruited via physician referrals, patient groups, or market research panels.  HCPs recruited from market research panels. | 72% (of total sample of patients) | Mean age of patients: 64.6 (±9.0) | IPF | Mixed method  Interview (in person and telephone).  Online survey. | This study identified several barriers to antifibrotic treatment, principally reflecting the differing views and values of patient and physician were identified in this study suggesting a need for better patient -physician communication about pharmacological therapy for IPF. |
| 22)Sampson  2015  UK | This study addresses the care needs of patients and carers at different stages of the IPF disease trajectory. | 48  Multi-site | Patients diagnosed with IPF and paired carers. | Two UK specialist interstitial lung disease clinics | Patients: 18 males  Carers: 6 males | Age range across the disease extent categories 56-87 | Patients with IPF(n=27)  Paired carers (n=21) | Cross sectional mixed methods study including interviews. | Patients diagnosed with IPF have a clear understanding of their prognosis but little understanding of how their disease will progress and how it will be managed.  Patients and carers outlined key elements of MDT activity capable of having significant impact on the care experience. They were structured around: focus on clinical encounters; timely identification of changes in health status and functional activity.; understanding of symptoms and medical interventions and coping strategies and carer roles. |
| 23)Tikellis  2020  Australia | Report the experiences of people who participated in the Peer Connect Service and to document the resources required to support such a service. | 32  Multi-site | Patients diagnosed with pulmonary fibrosis. | The Peer Connect Service managed by LFA, a national organization that provides support to people with a lung disease in Australia. | 56% | Mean ±SD range  71± 7 (53–89) | 32 peers consented to be interviewed. | Mixed methods study involving qualitative semi- structured interviews with Participants. | Major themes included the value of shared experiences, providing mutual support and the importance of shared personal characteristics (e.g. gender and hobbies) in allowing information and emotional support needs to be met. |
| 24)Tikellis  2022  Australia | Examine the organization of IPF care across Australia, how it aligns with guidance for best practice, and identified barriers and facilitators to best care. | 39  Multi-site | Purposive sampling of respiratory Physicians. | Hospitals and health networks with respiratory medicine services was identified through various sources that include the Australian Institute of Health and Welfare, the TSANZ website and internet searches. | unknown | unknown | IPF | Mixed Methods  Online questionnaire and semi-structured telephone interviews. | Approaches to diagnosis, treatment and access to referral services were generally consistent with best practice guidance.  Barriers related to inadequate staffing, lack of a nurse coordinator, inadequate access to clinical trials and funding models.  Telehealth technologies were perceived as facilitators to best care. |
| **Qualitative** | | | | | | | | | |
| 25)Bonella  2016  Austria,  Belgium,  France,  Germany,  Ireland,  Italy,  Spain,  UK,  Netherlans. | The aim of this study was to gather perceptions from European patient advocacy groups regarding inequalities and unmet needs in IPF care, in order to develop a Patient Charter. | Patient advocacy group members interviewed from 11 PG’s (n-12)  Working group (n-16)  Multi-site | Healthcare professionals and patient advocacy group members. | 11 Patient advocacy groups | unknown | unknown | IPF | Qualitative interviews  Individual telephone interview using a . | Five key themes were identified, the need for improved diagnosis, treatment access, holistic care, disease awareness and palliative care. |
| 26)Burnett  2019  Australia | The aim of this study was to assess the patient experience of modern IPF care. | 100  Registry  Multi-site | Patients diagnosed with IPF. | The Australian IPF Registry, | 61%  (total sample) | Age range 57-90 | IPF (N-100) | A Qualitative study using semi-structured interviews.  (telephone).  Grounded theory. | Dissatisfaction with information received particularly at the time of diagnosis.  The burden of travel to specialist centres and costs of treatment were significant. |
| 27)Cassidy  2021  Ireland. | Explore the palliative care and future health and social care planning requirements of individuals and families living with fibrotic interstitial lung disease (F-ILD) in Ireland. | 60 delegates attended with 39 stakeholders participating in discussion.  Single site | 39 stakeholders participating in discussion: patients (n=12), caregivers (n=13), healthcare and adjunct professionals (n=9), industry representatives (n=4) and clergy member (n=1). | World Café setting. | Patients (n=5 male), caregivers (n=6 male) | Unknown | F-ILD | World-Café qualitative research approach using a constructivist paradigm. | Palliative care is fundamental to the care and treatment of F-ILD’s, regardless of disease progression.  Unmet palliative care needs were identified as psychological and social support, disease education, inclusion of caregivers and practical and legal advice for disease progression and end-of-life planning. |
| 28)Delameillieure  2021  Belgium | To understand the care trajectory for patients with IPF based on the perspectives of patients and healthcare professionals. | 18  Single site | Healthcare professionals (n-9).  Patients (n-9) | IPF disease management program at UZ Leuven. | Patient group (67%) and caregiver group (40%) | Mean age 70 years for patient group | Patient group (n-9) IPF | Qualitative study using interviews. | Overall findings identified pitfalls and suggestions for improvement covering all elements of the chronic care model (CCM), primarily at the level of the individual patient and the care team.  Self-management support.  Patient reported outcomes and eHealth tools. |
| 29)Duck  2015  United Kingdom | To understand the perceptions, needs and experiences of patients with Idiopathic Pulmonary Fibrosis. | Patients (n-17)  Single site | Patients with a multidisciplinary team confirmed diagnosis of Idiopathic Pulmonary Fibrosis. | University Hospital of South Manchester Foundation Trust. | 7 Males | Median age 67 years | IPF (N=17) | Qualitative study.  Semi-Structured Interviews. | Three main themes were identified: ‘Struggling to get a diagnosis’; ‘Loss of the life I previously had’; and ‘Living with Idiopathic Pulmonary Fibrosis’. Patients reported struggling to get a diagnosis and coping with a life-limiting, rapidly progressive illness with no good treatment and few support structures. |
| 30)Giot  2012  Switzerland  Germany, France, Italy, Spain and the UK. | Researchers conducted a European survey of diagnosed IPF patients to identify unmet needs in the management of patients diagnosed with IPF and opportunities to improve care. | Patients (n=45)  Carers (n=18)  Multi-site | Patients diagnosed with IPF and in some interview’s carers were also interviewed. | European wide survey of patients in Germany, France, Italy, Spain and the UK. | Unknown | Median age of patient’s was 67 years. | IPF (n=45) | Qualitative study utilizing structured interviews. | In 58% of cases, diagnosis was protracted due to dismissal of symptoms and misdiagnosis.  Patients treated in specialist centres reported better satisfaction with care than those treated by generalists.  The patients cited negative impacts of IPF on almost all aspects of their lives, which could lead to depression. |
| 31)Kalluri  2021  Canada | To explore the perspectives, experiences and needs of patients with IPF, family caregivers, and healthcare professionals on ACP-related experiences to understand and inform a framework for advance care planning. | 20  Multi-site | HCP (N=10)  Patients (n=5) and caregivers (n=5). | Patients with IPF and family caregivers (PFCs) were recruited through the local Pulmonary Fibrosis Association. HCPs were recruited through email invitation letters to home care (HC) and acute care (AC). | Patients 60%  Caregivers 20% | Median age range patients: 69 (58-78)  Caregivers: 65 (20-73) | IPF (N=5) | Qualitative study using semi-structured interviews. | Participant perceptions included insufficient information and conversations occur late. Recommendations were to have earlier conversations; have open conversations; provide detailed information; and plan for end-of-life.  Professionals related delayed timing to poor end-of-life care and distressing deaths.  Acute care professionals perceived lack of clarity of roles and described personal, patient and caregiver distress. |
| 32)Lindell  2017  United States | Exploration of the perception of palliative care (PC) needs in patients with Idiopathic Pulmonary Fibrosis (IPF) and their family caregivers. | 13  Single site | Convenience sample of patients (n=5) and family caregivers (n=5) & family caregivers of decedent IPF patients (n=3). | The University of Pittsburgh Dorothy P. and Richard P. Simmons Centre for Interstitial Lung Disease at UPMC, a multidisciplinary centre for ILD treatment. | Male patients 100% | Mean age of patient with IPF 71.4 ± 7.2 | Time since diagnosis of IPF range 1-11 years | Qualitative study of patients and family caregivers using thematic analysis of focus group. | Four themes described 1) frustration with the diagnostic process and education received 2) overwhelming symptom burden, 3) hesitance to engage in advance care planning, and 4) comfort in receiving care from pulmonary speciality centre because of resources. |
| 33)Masefield  2019  UK  Ireland  Belgium  Italy | The aim of this study was to identify the communication challenges reported by patients and carers across Europe and elicit ideas for improvement. | 58  Multi-Site | Focus group participants: five patients and three carers in England; seven patients and four carers in Italy (two focus groups); six patients in Belgium; and twenty-three patients and ten carers in Ireland (five focus groups). | Four national patient organisations in Europe held focus groups: | Unknown | Unknown | IPF | Qualitative focus group study. | Patients and carers in Europe have unmet communication needs, which could be met by specialist physicians and specialist centres providing more effective information and signposting to support services, including support groups and patient organisations. |
| 34)Meadows  2017  USA | This study aimed to develop a new IPF patient/provider communication and disease education program. | 16  Single site | Patients diagnosed with IPF (n-12)  Caregivers (n=4). | The National Jewish Health (NJH) Interstitial Lung Disease (ILD) program | Patients (n=8 males)  Caregivers (n=2 males) | Unknown | IPF (n=12) | Qualitative focus groups. | Patents desired information on improving their own quality of life and caregivers desired information on acquiring knowledge and skills they felt necessary to improve their efficacy as caregivers of a patient with IPF.  Patient and caregiver directed information should come in multiple formats. |
| 35)Overgaard  2016  Denmark | The aim of this study was to increase knowledge of life with IPF for patients and family caregivers. | 49  Multi-site. | Patients (n=25)  Family Carers (n=24) | Specialist clinics at two University Hospitals | 15 male participants with IPF | Mean age Patients 71.1 (50-91) | IPF | Qualitative descriptive design using in-depth dyadic interviews with patients with IPF and family caregivers. | The following six themes were developed: information and disclosure, reactional dyssynchrony, perpetual vigilance, emotional ambivalence, gradual and tacit role shift, and adapted coping strategies. |
| 36)Ramadurai  2018  USA | This study sought to improve understanding of the informational needs of patients and their caregivers. | 17  Single site. | Patients diagnosed with IPF (n=13)  Caregivers (n=4) | Interstitial Lung Disease Programme at National Jewish Health | Patients 54%  Caregivers 50% | Mean age patients 68.1 (±7.2)  Caregivers 63.3 (±7.7) | IPF | Qualitative study using focus groups. | Patients wanted information on how to live well despite having IPF, practical information on how they could remain active and travel and how they could preserve their quality of life despite living with a life-threatening disease like IPF.  Caregivers wanted information on the general aspects of IPF. |
| 37)Russell  2016  UK  Italy  Germany | The aim of survey was to probe the impact of IPF on patients’ quality of life; the role of healthcare professionals and caregivers; the information needs of both patients and their caregivers; and patients’ perceptions of pirfenidone as a new treatment option for IPF. | 45  Multi-site. | Multidisciplinary team-confirmed diagnosis of IPF with disease duration >3 months. Patient with IPF (n=45) | Patients were invited to enrol through patient support groups (UK), specialist centres (Italy) or an advocacy group (Germany). | Male: 71% | Mean age 68.5 | IPF | Semi-structured, qualitative, in-depth patient interviews of 1-hour duration. | 87 % of patients reported that diagnosis took >1 year.  Patients reported that IPF had a significant physical and emotional impact on their quality of life.  The beneficial role played by caregivers and interstitial lung disease specialist nurses (where available) was specifically highlighted.  Information was often of poor quality, out of date, or in English only.  Patients’ perceptions of pirfenidone were largely positive and associated with ‘hope’ but were also influenced by the level of side effects experienced. |
| 38)Schoenheit  2011  Germany, France, Italy, Spain, UK | To gain further insights regarding patients’ perspectives on the diagnostic process, disease education, emotional well-being, and quality of life. | 45  Multi-site. | Patients (20% from each country) with physician confirmed diagnosis of IPF.  Caregivers participated in 18 (40%) of interviews. | Germany, France, Italy, Spain, UK | 49 % of patient sample | Median age: 67 | IPF | Qualitative In-depth interview (in person). | The median reported time from initial presentation to confirmed diagnosis of IPF was 1.5 years (range 1 year between initial presentation and a confirmed diagnosis of IPF).  The most common unmet needs cited by participants were disease education resources, access to centres of excellence, and familial support programs. |
| **Literature Reviews** | | | | | | | | | |
| 39)Bajwah  2017  United Kingdom | This review presents evidence for the epidemiology and management of the main symptoms of ILD and includes psychological needs and palliative care. | N/A | PF-ILD  ILD | N/A | N/A | N/A | ILD | Literature Review. | People living with ILD often suffer unmet physical and psychological needs throughout the disease journey.  Recent recommendations from the National Institute of Clinical Excellence promote the use of a new palliative care needs assessment tool. |
| 40)Chaaban  2021  USA | Literature review to highlight the palliative care needs of patients with ILD. | N/A | ILD | N/A | N/A | N/A | ILD | Literature Review. | Palliative care addresses symptoms in diseases where cure is unlikely or impossible, especially chronic diseases.  Palliative care can, and should, be implemented early to addresses goals of patients and caregivers. |
| 41)Kalluri  2019  Canada | This review sought to summarize the care pathway from the patient’s perspective, identifying current gaps in care, education, support and communication among patients with IPF, their caregivers, and care teams during the patient journey. | N/A | N/A | N/A | N/A | N/A | IPF | Literature review. | The ability to assess and control symptoms; develop self-efficacy; access to psychosocial, emotional, and spiritual support; and information on the disease and its course are important unmet needs for both patients with idiopathic pulmonary fibrosis and caregivers.  There is a huge need to support the multidisciplinary work focused on identifying gaps across the patient journey and PREMS and PROMs can be instrumental to this goal. |
| 42)Lee  2020  Australia | The supportive care needs of people with pulmonary fibrosis and their caregivers: A systematic review. | N/A | N/A | N/A | N/A | N/A | PF | Systematic Review. | A total of 35 studies were included in this review. The most frequently reported needs were in the domain of information/education and psychosocial/emotional needs  An additional domain ‘access to care’ was identified, this included access to peer support, psychological support, specialist centres and support for families of people with pulmonary fibrosis |
| 43)Robalo-Cordeiro  2017  Portugal | Manuscript preparation which was the outcome of a multidisciplinary meeting between pulmonology, radiology, and pathology clinicians on the use of antifibrotic agents in IPF. | N/A | N/A | N/A | N/A | N/A | N/A | Review. | Based on the available data from clinical trials and extension studies, the authors concluded that both Pirfenidone and Nintedanib have a significant effect on FVC decline.    Since IPF is a progressive lung disease, early diagnosis and treatment are crucial for slowing functional decline, reducing symptoms, and improving quality of life. |
| 44) Van Manen  2017  Netherlands | Literature review to summarise the evidence regarding quality of life for patients with IPF and discusses challenges in the management of this devastating disease. | N/A | N/A | N/A | N/A | N/A | IPF | Literature Review. | The review summarises the most recent insights into measuring and improving quality of life for patients with IPF.  The authors propose a new model for continuous care in IPF – the ‘ABCDE’ of IPF care. |
| **Guideline/Policy document** | | | | | | | | | |
| 45)EU-IPFF  Report  2020  UK | EU-IPFF benchmarking report on access to Idiopathic Pulmonary Fibrosis (IPF) care in Europe | 19 patient organisations involved in the study | Patient representatives and healthcare practitioners. | EU-IPFF member organisations and partners were invited to take part. There were 19 patient organisations from 17 EU Member States. | Unknown | Unknown | IPF | Qualitative study utilising phone interviews. | The Benchmarking Report outlines the current state of IPF care and management in Europe. It identifies best-performing countries along with challenges that require greater political attention and an immediate response. |
| 46)HSE  2020  Ireland | National Framework for the Integrated Care Programme for the Prevention and Management of Chronic Disease in Ireland | N/A | N/A | N/A | N/A | N/A | Chronic disease | HSE National Framework. | This report sets out the national framework which will describe an integrated approach to the prevention and management of chronic disease in Ireland over the coming years (2020-2025). |
| 47)Nice guideline  2017  United Kingdom | Idiopathic pulmonary fibrosis in adults: diagnosis and management | N/A | N/A | N/A | N/A | N/A | IPF | Guideline Document. | This guideline contains recommendations on the diagnosis of idiopathic pulmonary fibrosis and delivery of care to people with idiopathic pulmonary fibrosis. |
| 48)The Irish Thoracic Society  2018  Ireland | Irish Thoracic Society Position Statement on the Management of idiopathic pulmonary fibrosis. | N/A | N/A | N/A | N/A | 4/A | IPF | Position statement. | The standards of diagnosis and management of IPF need to be radically improved as outlined in the position statement.  All patients with IPF should have access to specialist centres and all appropriate treatments without undue delay |
| 49)The Irish Thoracic Society  2018  Ireland | Respiratory Health of the Nation 2018. | N/A | N/A | N/A | N/A | N/A | Respiratory conditions | Review of Respiratory conditions in Ireland. | Future research direction: Research needs to be focused on people and patients to identify barriers in terms of diagnosis, self-care and access to health services.  Research is also needed to focus on quality-of-life outcomes as well as evaluation focused on implementation of interventions. |
| 50)Irish Lung Fibrosis Association  2015  Ireland | National Patient Charter for Idiopathic Pulmonary Fibrosis | N/A | N/A | N/A | N/A | N/A | IPF | Patient Charter. | Six key patient entitlements outlined:  Early and accurate diagnosis.  Clear information about IPF.  Access to appropriate medicines and oxygen therapy.  Access to pulmonary rehabilitation and exercise programmes.  Early referral to the National Lung Transplant Unit for lung transplant assessment.  Access to psychological and palliative care services. |
| 51)Matthews  2018  Ireland | Patient charter on dying, death and bereavement in Ireland. | N/A | N/A | N/A | N/A | N/A | N/A | Patient Charter. | The importance of reforming health and social care systems and wider societal structures to ensure that dying people have access to appropriate care and psychological and spiritual supports is being increasingly recognised. |
| 52)Raghu    2022  United States | Idiopathic Pulmonary Fibrosis (an Update) and Progressive Pulmonary Fibrosis in Adults. An Official ATS/ERS/JRS/ALAT Clinical Practice Guideline | N/A | N/A | N/A | N/A | N/A | IPF | Clinical Practice Guidelines. | Recommendations for future research include the following: optimize strategies for addressing quality of life, including treatment of comorbidities, physical activity, emotional well-being, and palliation of symptoms. |
